# The effect of bivalent HPV vaccination against invasive cervical cancer and CIN3+ in the Netherlands: a national linkage study

**DOI:** 10.1101/2025.02.27.25322519

**Authors:** Marit Middeldorp, Jesca G.M. Brouwer, Janneke W. Duijster, Mirjam J. Knol, Folkert J. van Kemenade, Albert G. Siebers, Johannes Berkhof, Hester E. de Melker

## Abstract

**Background:** The protective effect of HPV vaccination against cervical cancer has been demonstrated in registry linkage studies. The start age of screening in those studies was lower than 25 years. We estimated the vaccine effectiveness of bivalent HPV16/18 vaccination against invasive cervical cancer and cervical intraepithelial neoplasia grade 3 (CIN3+) in the Netherlands where routine screening starts at age 30 years.

**Methods:** We linked the vaccination status of women born in year 1993 who were eligible for HPV vaccination at age 16 years with histopathological results recorded in the nationwide pathology databank (Palga). Cumulative risks of invasive cervical cancer and CIN3+ were estimated for fully vaccinated (3 doses or 2 doses ≥150 days apart), partially vaccinated, and unvaccinated women. Vaccine effectiveness estimates were adjusted for differences in screening participation between the vaccine groups.

**Findings:** A total of 103,059 women were included, of whom 47,130 were fully vaccinated, 5,098 were partially vaccinated, and 50,831 were unvaccinated. Five (0·011%) cancers were observed in fully vaccinated, two (0·039%) in partially vaccinated, and 42 (0·083%) in unvaccinated women. The vaccine effectiveness in fully vaccinated women was 91·5% (95% CI 78·9, 96·6) against cancer and 81·2% (95% CI 78·4, 83·7) against CIN3+. The vaccine effectiveness in partially vaccinated women was 48·1% (95% CI -56·8, 82·8) against cancer and 58·4% (95% CI 45·3, 68·3) against CIN3+.

**Interpretation:** The high effectiveness of bivalent HPV vaccination against cervical cancer and CIN3+ and the low cancer incidence supports a screening start age of 30 years in vaccinated women.

**Research in context:** *Evidence before this study:* We searched Pubmed and Google Scholar with the search terms (“Cervical Cancer”) AND (“HPV” OR “human papillomavirus”) AND (“vaccination”). Articles published in English were searched until January 2, 2025. Studies from Sweden, Denmark, and Scotland were identified linking individual vaccination, screening and cancer registry data. The start age of screening in these studies was 23-25 years. They showed a strong effectiveness in preventing cervical cancer following the introduction of bivalent and quadrivalent HPV vaccination.

*Added value of this study:* We observed a very low absolute incidence of cervical cancer in vaccinated women and a much lower incidence of cervical cancer and CIN3+ in women vaccinated at age 16 compared with unvaccinated women, in a setting where routine screening starts at age 30. By linking the vaccination registry to the nationwide pathology databank, we were able to adjust for screening non-attendance in the incidence of cancer and CIN3+ over a 15 year period.

*Implications of all the available evidence:* Our study supports a start age of screening of at least 30 years in women vaccinated at a young age. Avoiding screening before age 30 in these women is expected to substantially reduce the harms associated with screening and treatment.

## Introduction

A persistent human papillomavirus (HPV) infection is the established cause of invasive cervical cancer^1^, being the fourth-most common cancer in women worldwide.^2^ To eliminate cervical cancer, the importance of HPV vaccination in combination with routine cervical cancer screening and appropriate treatment has been emphasized by the World Health Organization (WHO).^3^

In the Netherlands, bivalent HPV16/18 vaccination with a 3-dose schedule for girls age 12-13 years was implemented in the National Immunisation Programme (NIP) in 2010. The implementation was preceded by a catch-up campaign in 2009, in which bivalent vaccination was offered to 13-16-year olds girls (i.e. born between 1993 and 1996). Since its introduction in the Netherlands, the coverage of the HPV vaccination program has been relatively low, ranging between 46% and 63%.^4^ In addition to vaccination, women aged 30 to 60 years are invited for routine cervical cancer screening every 5 to 10 years, where the screening interval depends on the women’s age and previous test result.^5^ Primary screening options include a high-risk (hr) HPV test that can be conducted on either a home-collected sample or a physician-collected sample. Furthermore, some women have a physician-collected sample on medical indication prior to the start age of routine screening.^6^ In 2023, women eligible for catch-up HPV vaccination at age 16 (i.e. born in year 1993) were invited to participate in routine cervical cancer screening at age 30. This allows us to assess the real-world vaccine effectiveness of the bivalent HPV vaccine against invasive cervical cancer.

In this study, we estimated the vaccine effectiveness (VE) of bivalent HPV vaccination against invasive cervical cancer and cervical intraepithelial neoplasia grade 3 or worse (CIN3+) in the Netherlands. The results presented are the first from a nationwide cohort vaccinated at age 16 and eligible for routine screening at age 30. This setting is also relevant for countries where screening starts before the age of 30 and a later starting age of screening is being considered in order to retain a programme where the benefits outweigh the harms. To date, only Italy has increased the start age of screening for vaccinated women from 25 to 30 years.^7^ Most other countries are awaiting data on the longevity of vaccine protection before increasing the starting age. Our study can inform local decision-making by providing further evidence on the long-term residual risk of cancer in vaccinated women.

## Methods

### Study population and linkage

For this national linkage study, we selected all women registered in the national vaccination registry of the Netherlands (Praeventis) with birth year 1993 (n=109,227). Praeventis is the administrative database from the NIP that includes all children registered as residents in the Netherlands and tracks all vaccines administered within the NIP. The selected women were linked to results of cervical samples and/or tissue sampling recorded in the Dutch nationwide pathology databank (Palga).

Deterministic linkage was performed using surname, birthdate, sex, and, if necessary, the first letter of the given name. Women were included in the statistical analyses if they were alive and living in the Netherlands at any moment in 2009 (n=104,661). Women were excluded when vaccinated in a calendar year other than 2009 or 2010, or with an HPV vaccine other than the bivalent vaccine (n=1,602).

Institutional Review Board (IRB) of the Centre for Clinical Expertise at the National Institute for Public Health and the Environment (RIVM), the Netherlands, waived ethical approval for this work. Approval of the study was obtained from the national vaccination registry of the Netherlands (Praeventis) and the nationwide pathology databank of the Netherlands (Palga).

### Vaccination, screening, and histopathological data

Praeventis contains all relevant information on the HPV vaccination status of each individual in the Netherlands. Women who received the bivalent vaccine were considered fully vaccinated if they received three doses with an interval of 21 to 150 days between dose 1 and 2 and an interval of at least 120 days between dose 2 and 3, or if two doses were administered at least 150 days apart. They were classified as partially vaccinated if they received one dose, or if they received two or three doses in a schedule that violated the criteria for being fully vaccinated. Women who did not receive any dose were classified as unvaccinated.

Palga contains hrHPV- and cytological test results of cervical screening samples and histological results of cervical tissue samples. Cervical tissue samples may be collected after an hrHPV-positive result in the cervical cancer screening programme, where women can choose between self-collection or physician-collection. Cervical tissue samples may also be collected outside the screening programme, based on gynaecological complaints. We included cytology results up to February 15, 2024 and histological follow-up after cytology until April 1, 2024. This time window provided sufficient time to obtain a histological confirmation after cytological high-grade squamous intraepithelial lesion (HSIL) as 90% of the women with cytological HSIL had a histological result within 1·5 months. hrHPV testing results were collected until 1 April 2024.

### Statistical analyses

Descriptive analyses were performed for fully vaccinated, partially vaccinated, and unvaccinated women. We calculated the proportion of women with available test results (hrHPV, cytological, and/or histological result) both within and/or outside the screening program, the proportion of women choosing self-collection, and the proportion of HPV-positive test results among women in the screening program. Furthermore, we reported the number of histologically confirmed cervical cancers, CIN3, and CIN2.

Cumulative risks of cervical cancer and CIN3+ were calculated for each vaccine group (fully vaccinated, partially vaccinated and unvaccinated). Women were included in the denominator of the risk estimate, irrespective of whether they had a result in Palga. Cumulative risks for end-points cervical cancer and CIN3+ were also calculated for each vaccine group by age of diagnosis (<25 years, 25-29 years, and >=30 years). Jeffrey’s intervals were calculated around these risks.^8^

We calculated the adjusted cumulative cancer (and CIN3+) risk per vaccination group by summing the cumulative risk detected outside the screening program and the cumulative risk detected within the screening program, where we divided the latter risk by the screening participation percentage.

Women with CIN grade 3 or worse detected outside the screening program were excluded from the denominator when calculating the cumulative risk detected within the screening program. The cumulative risk ratios (Cumulative RRs, CRRs) were calculated as the cumulative risk in either fully or partially vaccinated women divided by the cumulative risk in unvaccinated women. The VE estimates were calculated as *(1 - CRR) * 100%*. Estimates were accompanied by 95% confidence intervals (95% CIs).

Crude VE estimates were additionally stratified by socioeconomic status (SES) based on a woman’s 4-digit postal code available in Praeventis. This measure of SES is a summary score which reflects financial wealth, education level, and recent labour participation at average household level in 2021 for each postal code (i.e. neighbourhood).^9^ The VE estimates stratified by SES were pooled using the Mantel-Haenszel method.

A sensitivity analysis was performed with a less strict approach regarding vaccination status. This analysis additionally included women who were vaccinated in calendar years other than 2009 or 2010 or who received an HPV vaccine other than the bivalent vaccine (n=1,602).

All analyses were performed using R (version 4.4.0).

## Results

### Characteristics of the study population

A total of 103,059 women were included in the analyses. Of these, 47,130 (45·7%) were fully vaccinated with either two doses (2·0%) or three doses (98·0%), 5,098 (5·0%) were partially vaccinated, and 50,831 (49·3%) were unvaccinated (Table 1). The percentage of women living in a neighbourhood with a low SES was lowest for fully vaccinated women (49·5%) and highest for partially vaccinated women (58·7%). A higher participation rate in the routine cervical cancer screening program was observed among fully vaccinated women (58·8%) than among partially vaccinated women (43·3%) and unvaccinated women (44·2%). The percentage of women with test results available outside the screening program was similar for fully vaccinated women and unvaccinated women (24·9% and 23·9%, respectively) and slightly higher for partially vaccinated women (28·5%). Additionally, 31·3% of the fully vaccinated women, 42·2% of the partially vaccinated women, and 44·7% of the unvaccinated women never had a cervical screening- or tissue sample.

**Table 1.** Characteristics of the study population.

|  | **Fully vaccinated** | **Partially vaccinated** | **Unvaccinated** |
| --- | --- | --- | --- |
| **Number of women** | 47,130 (45.7%) | 5,098 (5.0%) | 50,831 (49.3%) |
| **Number of doses** |  |  |  |
| 3 | 46,190 (98.0%) | 11 (0.22%) | 0 (0.0%) |
| 2 | 940 (2.0%) | 2,837 (55.6%) | 0 (0.0%) |
| 1 | 0 (0.0%) | 2,250 (44.1%) | 0 (0.0%) |
| 0 | 0 (0.0%) | 0 (0.0%) | 50,831 (100.0%) |
| **Neighbourhood socioeconomic status** |  |  |  |
| High | 9,391 (19.9%) | 749 (14.7%) | 9,248 (18.2%) |
| Medium | 14,277 (30.3%) | 1,338 (26.2%) | 14,609 (28.8%) |
| Low | 23,322 (49.5%) | 2,992 (58.7%) | 26,813 (52.8%) |
| Unknown | 140 (0.30%) | 19 (0.37%) | 161 (0.32%) |
| **Cervical cancer screening participation rate** | 27,692 (58.8%) | 2,207 (43.3%) | 22,478 (44.2%) |
| **Origin of cervical sample** |  |  |  |
| Within screening program only | 20,618 (43.7%) | 1,492 (29.3%) | 15,964 (31.4%) |
| Outside screening program only | 4,672 (9.9%) | 740 (14.5%) | 5,652 (11.1%) |
| Both within and outside screening program | 7,074 (15.0%) | 715 (14.0%) | 6,514 (12.8%) |
| No cervical sample | 14,766 (31.3%) | 2,151 (42.2%) | 22,701 (44.7%) |
| **Participation method in screening program** |  |  |  |
| Self-sampling kit at home | 16,492 (59.6%) | 1,323 (59.9%) | 13,361 (59.4%) |
| Smear at the general practitioner | 11,200 (40.4%) | 884 (40.1%) | 9,117 (40.6%) |
| **HPV genotyping result within screening program** |  |  |  |
| 16\|18 | 30 (0.11%) | 12 (0.54%) | 821 (3.7%) |
| 16\|18 and other hrHPV type* | 17 (0.06%) | 8 (0.36%) | 763 (3.4%) |
| Other hrHPV type* | 3,977 (14.4%) | 354 (16.0%) | 2,773 (12.3%) |
| hrHPV negative | 23,272 (84.0%) | 1,781 (80.7%) | 17,684 (78.7%) |
| HPV genotyping result unknown** | 370 (1.3%) | 50 (2.3%) | 441 (1.8%) |
| hrHPV status unknown | 26 (0.09%) | 2 (0.09%) | 26 (0.12%) |
| **HPV genotyping result outside screening program** |  |  |  |
| 16\|18 | 8 (0.07%) | 3 (0.21%) | 354 (2.9%) |
| 16\|18 and other hrHPV type* | 6 (0.05%) | 4 (0.27%) | 380 (3.1%) |
| Other hrHPV type* | 629 (5.4%) | 80 (5.5%) | 582 (4.8%) |
| HrHPV negative | 2,861 (24.4%) | 337 (23.2%) | 2,774 (22.8%) |
| HPV genotyping result unknown** | 1,084 (9.2%) | 181 (12.4%) | 1,562 (12.8%) |
| hrHPV status unknown | 7,158 (60.9%) | 850 (58.4%) | 6,514 (53.5%) |
| **Number of cervical cancers** |  |  |  |
| Within screening program | 0 (0.0%) | 1 (50.0%) | 17 (40.5%) |
| Outside screening program | 5 (100.0%) | 1 (50.0%) | 25 (59.5%) |
| Total | 5 | 2 | 42 |
| **Number of CIN3** |  |  |  |
| Within screening program | 71 (42.3%) | 15 (50.0%) | 348 (45.7%) |
| Outside screening program | 95 (57.2%) | 15 (50.0%) | 413 (54.3%) |
| Total | 166 | 30 | 761 |
| **Number of CIN2** |  |  |  |
| Within screening program | 90 (30.6%) | 8 (16.0%) | 234 (35.9%) |
| Outside screening program | 204 (69.4) | 42 (84.0%) | 417 (64.1%) |
| Total | 294 | 50 | 651 |

Abbreviations: CIN3, cervical intraepithelial neoplasia grade 3. CIN2, cervical intraepithelial neoplasia grade 2. \*Other hrHPV types include HPV types 31, 33, 35, 39, 45, 51, 52, 56, 58, 59, 66 and 68. \*\* hrHPV positive women without hrHPV genotyping result.

For women who participated in the routine screening program, approximately 60% used a self-sampling kit at home, while 40% visited their general practitioner for a cervical scrape (Table 1). 84·0% of fully vaccinated women, 80·7% of partially vaccinated women, and 78·7% of unvaccinated women had a negative hrHPV test in the cervical screening programme. HPV16/18 positivity was highest in unvaccinated women, but also higher in partially vaccinated women than in fully vaccinated women.

The majority (57·6%) of women with a cervical sample collected outside the screening program did not undergo hrHPV testing.

### Cervical cancer and cervical intraepithelial neoplasia

#### Cumulative risks and vaccine effectiveness

In total, 49 women were diagnosed with cervical cancer. Of these, five were fully vaccinated (cumulative risk: 0·011%), two were partially vaccinated (0·039%), and 42 were unvaccinated (0·083%) (Table 1 and 2). The five cervical cancers diagnosed in fully vaccinated women were all identified outside the screening program. In partially vaccinated women, one cancer was diagnosed in the screening program and one outside the screening program. In unvaccinated women, 17 (40·5%) of the cancers were detected in the screening program and 25 (59·5%) were detected outside the screening program. Also within each age group at the time of diagnosis, unvaccinated women had higher risks of cervical cancer and CIN3+ compared to vaccinated women (Figure 1).

**Table 2.** Cumulative risks of cervical cancer and cervical intraepithelial neoplasia grade 3 or worse (CIN3+) by vaccination status with corresponding vaccine effectiveness estimates.

|  | Number of<br>cervical<br>cancer | Risk, %<br>(95% CI) | Crude VE<br>(95% CI) | Adjusted* VE<br>(95% CI) | Number of<br>CIN3+ | Risk, %<br>(95% CI) | Crude VE<br>(95% CI) | Adjusted* VE<br>(95% CI) |
| --- | --- | --- | --- | --- | --- | --- | --- | --- |
| Fully vaccinated<br>(N=47,130) | 5 | 0.011<br>(0.0045,0.025) | 87.2<br>(67.6, 94.9) | 91.5<br>(78.9, 96.6) | 171 | 0.36<br>(0.31, 0.42) | 77.0<br>(72.9, 80.5) | 81.2<br>(78.4, 83.7) |
| Partially vaccinated<br>(N=5,098) | 2 | 0.039<br>(0.011, 0.14) | 52.5<br>(-96.1, 88.5) | 48.1<br>(-56.8, 82.8) | 32 | 0.63<br>(0.45, 0.89) | 60.3<br>(43.5, 72.1) | 58.4<br>(45.3, 68.3) |
| Unvaccinated<br>(N=50,831) | 42 | 0.083<br>(0.061 , 0.11) | Ref. | Ref. | 803 | 1.58<br>(1.48, 1.69) | Ref. | Ref. |

Abbreviations, N: Number of women; CIN3+: cervical intraepithelial neoplasia grade 3 or worse; VE: vaccine effectiveness; Risk: Crude cumulative risk; CI: confidence interval; Ref.: reference category. \*Adjusted for differences in cervical cancer screening participation by vaccination status.

**Figure 1.**
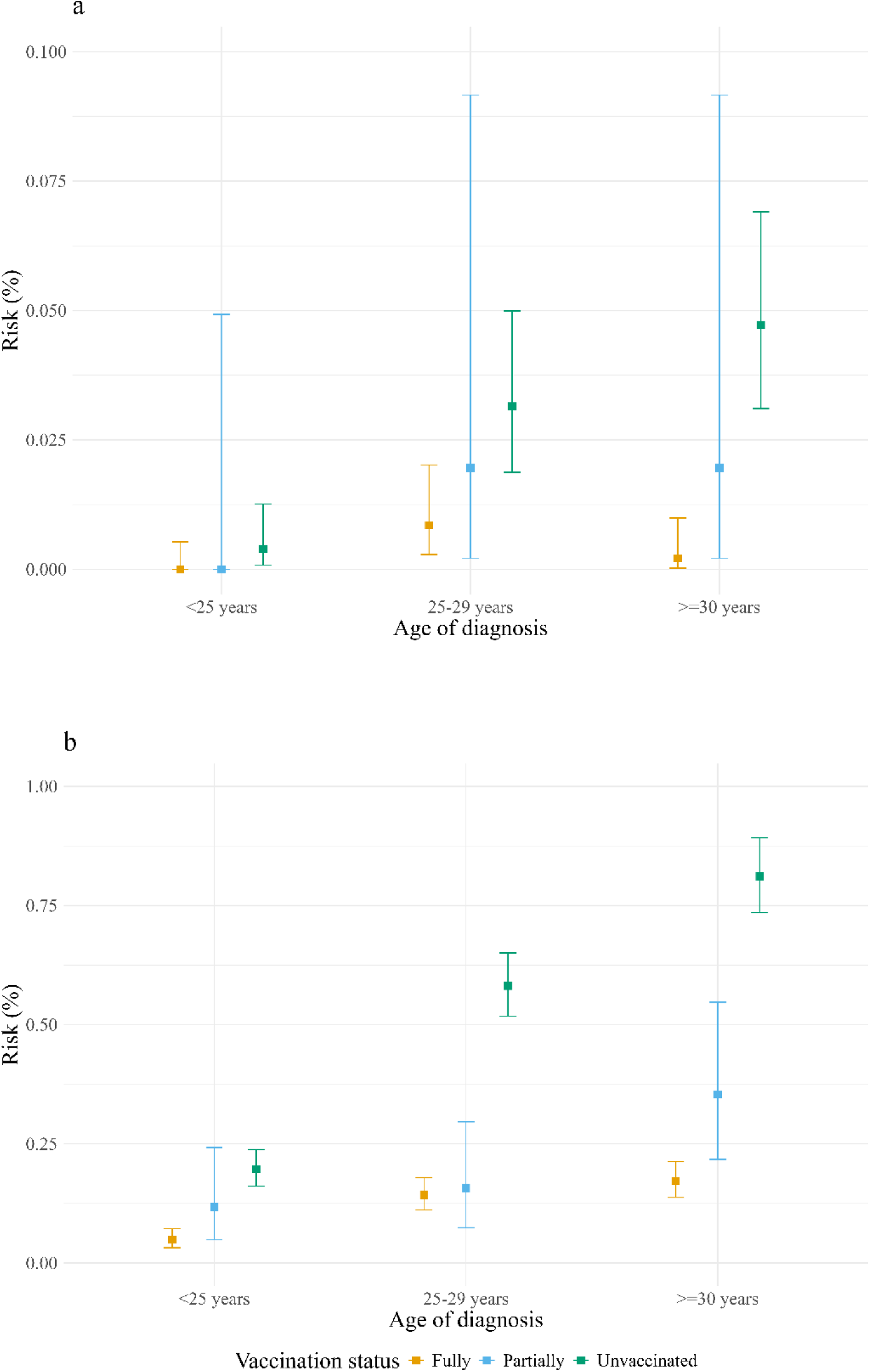
Cervical cancer (panel a) and CIN3+ (panel b) risk by vaccination status and age of diagnosis. All risks are displayed with Jeffrey’s interval. CIN3+, cervical intraepithelial neoplasia grade 3 or worse. >=30 years, diagnosis at ages 30 or 31.

The VE for full vaccination at 16 years of age with the bivalent HPV vaccine was 87·2% (95% CI 67·6, 94·9) (Table 2). When accounting for the differences in screening participation, the VE was 91·5% (95% CI 78·9, 96·6). For partially vaccination, the adjusted VE against cervical cancer was 48·1% (95% CI -56·8, 82·8).

In total, 957 women were diagnosed with CIN3 (Table 1). Of them, 166 were fully vaccinated, 30 were partially vaccinated, and 761 were unvaccinated. The crude and adjusted effectiveness against CIN3+ for fully vaccinated women compared to unvaccinated women were 77·0% (95% CI 72·9, 80·5) and 81·2% (95% CI 78·4, 83·7), respectively (Table 2). The adjusted effectiveness against CIN3+ for partially vaccinated women compared to unvaccinated women was 58·4% (95% CI 45·3, 68·3).

Furthermore, 995 women were diagnosed with CIN2 (Table 1). Of these, 294 were fully vaccinated, 50 were partially vaccinated, and 651 were unvaccinated. Table 1 shows the number of CIN3 and CIN2 detected within and outside the screening program.

The crude VE estimates adjusted by neighbourhood SES demonstrated similar estimates for fully vaccinated women against cervical cancer (87·3%, 95% CI 67·9, 96·1) and CIN3+ (77·4, 95% CI 73·3, 81·0) when compared to the crude VE estimates from the main analyses reported in Table 2.

#### HPV genotyping results

In the screening program, one partially vaccinated woman diagnosed with cervical cancer was positive for HPV16/18 along with another hrHPV type (Table S1). Among unvaccinated women diagnosed with cervical cancer in the screening program, 9 (52·9%) were positive for HPV16/18 and 5 (29·4%) were positive for HPV16/18 along with another hrHPV type.

Among fully vaccinated women diagnosed with CIN3+ in the screening program, 3 (4·2%) were positive for at least HPV16 or HPV18, while 67 (94·4%) were positive for an hrHPV type other than HPV16 or HPV18 (Table S1). For partially vaccinated women, these numbers were 5 (31·3%) and 11 (68·8%), respectively. For unvaccinated women, these numbers were 284 (77·8%) and 76 (20·8%), respectively.

#### Sensitivity analysis

In the sensitivity analysis, all women who were eligible for HPV vaccination in 2009 were included (n=104,661). The VE estimates were similar to those in the main analysis for both cervical cancer and CIN3+ (Table S2). The adjusted VE against cervical cancer among fully vaccinated women compared to unvaccinated women was 91·8% (95% CI 79·5, 96·7). For CIN3+, the adjusted VE was estimated at 80·4% (95% CI 77·5, 82·9). For partially vaccinated women, the adjusted VE estimates were 50·7% (95% CI -49·5, 83·7) against cervical cancer and 59·8% (95% CI 47·2, 69·3) against CIN3+.

## Discussion

With this national linkage study we showed that bivalent HPV vaccination at 16 years of age is highly effective against invasive cervical cancer and CIN3+ when comparing fully vaccinated women to unvaccinated women. For partially vaccinated women, the VE against CIN3+ was lower than for fully vaccinated women and the VE against cancer was not significant, probably due to the small number of partially vaccinated women.

The observed crude effectiveness of 87% against cervical cancer is consistent with previous research from Scotland, which reported an effectiveness of 86% among women vaccinated at 14-16 years of age with the bivalent HPV vaccine.^10^ In Sweden and Denmark, comparable VE estimates were observed among women vaccinated with the quadrivalent vaccinate before 17 years of age.^11,12^ In England, an observational study without individual linkage between the vaccine and cancer registry reported a slightly lower effect (62%) among women who were offered bivalent vaccination at 14-16 years of age.^13^

The lower cervical cancer screening participation rate among unvaccinated women in our study has also been observed in other countries.^14-16^ A higher screening participation among vaccinated women increases the probability of detecting an asymptomatic cervical cancer in this group, and may lead to a negatively biased VE estimate.^12^ After accounting for the difference in screening participation rate by vaccination status, the VE against cervical cancer increased to 92% for fully vaccinated women in our study. Our finding of a slightly lower protective effect against CIN3+ compared to invasive cancer aligns with previous studies from Denmark^11,17^, and might be due to the stronger association between cervical cancer and HPV types 16 and 18 than between CIN3 and these HPV types.^18^ A high effectiveness against CIN3+ was still observed for fully vaccinated women and further increased after adjusting for differences in screening participation. In England and Sweden, the effect estimates against CIN3+ were similar as in our study.^13,19^ In Australia, the quadrivalent vaccine demonstrated a slightly lower VE (57%).^20^

In our study, we observed five cases of cervical cancer in women fully vaccinated at 16 years of age. Globally, HPV types 16 and 18 are responsible for approximately 75% of cervical cancer cases, and HPV types 31, 33, and 45 account for an additional 11% of cervical cancers.^21^ Consequently, even in the presence of cross-protection^22^, fully vaccinated women can still develop cervical cancer due to hrHPV types that are not targeted by the vaccine. This underscores that cervical cancer screening still offers health gains, also to those who are fully vaccinated, potentially through customized screening strategies for vaccinated women to prevent overtreatment.^23^

A considerable number of women in our study population did not make use of any of the cervical cancer prevention methods, i.e. vaccination and screening. These women are at increased risk of developing cervical cancer. They may still benefit from the indirect effects of HPV vaccination that we observed in the Netherlands^24^, although the herd effect is expected to be limited for the first vaccinated cohort and will become stronger for future cohorts. In addition, switching to gender-neutral vaccination may become beneficial for unvaccinated women as well since the expected herd effect is expected to increase.^23,25^

Cervical samples collected following a medical consultation, rather than as part of the screening program, creates the potential to interrupt the progression of cervical cancer when pre-cancerous lesions at a pre-screening age are detected and treated. Consequently, women with pathology results from medically initiated cervical samples may contribute to a lower incidence of cervical cancer. In our study, the percentage of women with a pathology results of a medically initiated cervical sample (i.e. outside the screening program) was comparable for vaccinated and unvaccinated women. Therefore, it is unlikely that the observed results are biased by a difference in screening outside the program between vaccinated and unvaccinated women.

We observed lower vaccine effectiveness in the partially vaccinated group compared with the fully vaccinated group. One possible explanation is that partially vaccinated women have a higher risk of acquiring HPV infection than fully vaccinated women. This effect may translate into lower effectiveness, as women in our population were vaccinated at the age of 16 years and pre-vaccination exposure to HPV cannot be excluded. Although we did not collect individual data on risk behaviour, partially vaccinated women had a lower cervical screening uptake, lower neighbourhood SES, higher HPV16/18 prevalence at routine screening, and a slightly higher HPV prevalence for other genotypes than fully vaccinated women. This may be associated with lower levels of health-protective or higher sexual risk behaviour.^26^ A second possible explanation is that completing the full vaccination schedule gives a better protection against cervical cancer and CIN3+ than partial vaccination. Our study is consistent with a recent Scottish linkage study which reported a slightly higher risk of cancer after partial vaccination with the bivalent vaccine than after full vaccination at age 14 years or older.^10^ A recent Swedish linkage study also observed comparable vaccine effectiveness against CIN2+ for one, two and three doses of the quadrivalent vaccine when given before the age of 15, but slightly higher risks for one dose compared to three doses in older age groups.^27^ In addition, two ongoing randomized controlled trials indicate that one dose provides stable high immunogenicity levels over five years of follow-up, although geometric mean concentration levels (GMCs) were slightly lower with a single dose than with two doses. A strong protection against HPV infections over three years of follow-up was observed.^28-32^ In summary, several studies suggest that a single-dose vaccination schedule is highly effective, but close monitoring of current and future vaccinated cohorts remains important to consider whether specific age groups might benefit from more than one dose to achieve optimal protection against cancer and CIN3.

The cohort included in this study consisted of women eligible for bivalent HPV vaccination at 16 years of age (catch-up vaccination). Consequently, it is possible that women were exposed to HPV before vaccination given that the median age of sexual debut among female adolescents in the Netherlands in 2012 is 16·8 years.^33^ Other studies found that a higher vaccine effectiveness against cervical cancer is associated with a younger age of HPV vaccination for both the bivalent and quadrivalent vaccine.^10-13^ Therefore, we may in the future observe an even higher vaccine effectiveness against cervical cancer in the Netherlands, as the age of invitation for HPV vaccination was lowered to 12 years in 2014 and further reduced to 9 years in 2022. Moreover, it has been suggested that the number of cervical cancer cases will decrease across the population due to more pronounced indirect effects among the unvaccinated population if more birth cohorts receive vaccination.^11,24^

An important strength of this study is that it includes women who enter cervical cancer screening at the age of 30, i.e. at a later age than has been reported in other countries. This makes our linkage between the national databases on vaccination, screening and cancer data a valuable resource for assessing the long-term effectiveness of vaccination and the benefits and harms of initiating screening at this age. Additionally, by using the information from the national vaccination registry, the risk of misclassification regarding the vaccination status was minimized. However, our study has some limitations. First, we were unable to adjust for variables that could potentially confound the association between HPV vaccination and the risk of cervical cancer such as sexual behaviour and exposure to HPV pre-vaccination. However, it is reassuring that we did not observe considerable differences in the positivity for other hrHPV types between fully vaccinated and unvaccinated women. We were able to account for SES, although not at individual level but at the 4-digit zip code level based on financial wealth, education level and recent labour participation at household level. After adjusting for neighbourhood SES, the estimates were comparable to the crude estimates from the main analyses. A third limitation is the dataset’s incompleteness regarding emigration records. This incompleteness could have affected the results in case of substantial differences in migration between vaccinated and unvaccinated women, although this is unlikely, as suggested by Schurink-van ‘T Klooster et al.^6^ A fourth limitation is that follow-up data are incomplete for hrHPV-positive women in the screening programme who were invited for repeat cytology after one year. This includes women with low grade abnormal cytology (ASCUS/LSIL) who are negative for HPV16/18 and women with normal cytology.^5^

In conclusion, we observed a high effectiveness of the bivalent HPV vaccination administered at 16 years of age. The extent of protection against cervical cancer for partially vaccinated women could not be determined with certainty due to the low number of cases. We will continue to monitor the effectiveness of HPV vaccination in this birth cohort and in subsequent cohorts. As more individuals who were eligible for HPV vaccination enter the cervical cancer screening program at age 30, we anticipate getting further confirmation of the long-term vaccine effectiveness in preventing cervical cancer and the building up of indirect protective effects by age.

## Data Availability

Aggregated data that support the findings of this study are available upon reasonable request from the corresponding author. The data are not publicly available since this could compromise the privacy of the participants.

