## Supplementary material for "The effect of bivalent HPV vaccination against invasive cervical cancer and CIN3+ in the Netherlands: a national linkage study"

**Table of contents**

Supplementary Table 1. Page 2

Supplementary Table 2. Page 3

**Table S1. HPV genotyping results for women with cervical cancer and cervical intraepithelial neoplasia grade 3 or worse (CIN3+).**

| | Cervical cancer | | CIN3+ | |
|---|---|---|---|---|
| **Diagnosed** | Within screening program, n = 18 | Outside screening program, n = 31 | Within screening program, n = 452 | Outside screening program, n = 554 |
| **HPV types** | | | | |
| **Fully vaccinated** | | | | |
| 16\|18 | 0 (0·0%) | 0 (0·0%) | 2 (2·8%) | 0 (0·0%) |
| 16\|18 and other hrHPV* | 0 (0·0%) | 0 (0·0%) | 1 (1·4%) | 0 (0·0%) |
| Other hrHPV* | 0 (0·0%) | 1 (20·0%) | 67 (94·4%) | 15 (15·0%) |
| hrHPV negative | 0 (0·0%) | 0 (0·0%) | 0 (0·0%) | 3 (3·0%) |
| HPV genotyping result unknown** | 0 (0·0%) | 1 (20·0%) | 1 (1·4%) | 55 (55·0%) |
| hrHPV status unknown | 0 (0·0%) | 3 (80·0%) | 0 (0·0%) | 27 (27·0%) |
| **Partially vaccinated** | | | | |
| 16\|18 | 0 (0·0%) | 0 (0·0%) | 3 (18·8%) | 0 (0·0%) |
| 16\|18 and other hrHPV* | 1 (100·0%) | 0 (0·0%) | 2 (12·5%) | 0 (0·0%) |
| Other hrHPV* | 0 (0·0%) | 0 (0·0%) | 11 (68·8%) | 2 (12·5%) |
| hrHPV negative | 0 (0·0%) | 0 (0·0%) | 0 (0·0%) | 1 (6·2%) |
| HPV genotyping result unknown** | 0 (0·0%) | 1 (100·0%) | 0 (0·0%) | 6 (37·5%) |
| hrHPV status unknown | 0 (0·0%) | 0 (0·0%) | 0 (0·0%) | 7 (43·8%) |
| **Unvaccinated** | | | | |
| 16\|18 | 9 (52·9%) | 4 (16·0%) | 157 (43·0%) | 37 (8·4%) |
| 16\|18 and other hrHPV* | 5 (29·4%) | 0 (0·0%) | 127 (34·8%) | 31 (7·1%) |
| Other hrHPV* | 2 (11·8%) | 2 (8·0%) | 76 (20·8%) | 26 (5·9%) |
| hrHPV negative | 0 (0·0%) | 0 (0·0%) | 0 (0·0%) | 1 (0·2%) |
| HPV genotyping result unknown** | 1 (5·9%) | 10 (40·0%) | 5 (1·4%) | 222 (50·7%) |
| hrHPV status unknown | 0 (0·0%) | 9 (36·0%) | 0 (0·0%) | 121 (27·6%) |

Abbreviations: CIN3+: cervical intraepithelial neoplasia grade 3 or worse. *Other hrHPV types include HPV types 31, 33, 35, 39, 45, 51, 52, 56, 58, 59, 66 and 68. ** HrHPV positive women without hrHPV genotyping result.

**Table S2**. **Cumulative risks of cervical cancer and cervical intraepithelial neoplasia grade 3 or worse (CIN3+) by vaccination status with corresponding vaccine effectiveness estimates.** Results from the sensitivity analysis in which women were also included when vaccinated in calendar years other than 2009/2010 or when vaccinated with another HPV vaccine than the bivalent vaccine.

| | **Number of cervical cancer** | **Risk, % (95% CI)** | **Crude VE (95% CI)** | **Adjusted VE (95% CI)** | **Number of CIN3+** | **Risk, % (95% CI)** | **Crude VE (95% CI)** | **Adjusted VE (95% CI)** |
|---|---|---|---|---|---|---|---|---|
| Fully vaccinated (N=48,501) | 5 | 0·010 (0·0044, 0·024) | 87·5 (68·5, 95·1) | 91·8 (79·5, 96·7) | 184 | 0·38 (0·33, 0·44) | 76·0 (71·8, 79·5) | 80·4 (77·5, 82·9) |
| Partially vaccinated (N=5,329) | 2 | 0·038 (0·010, 0·14) | 54·6 (-87·6, 89·0) | 50·7 (-49·5, 83·7) | 33 | 0·62 (0·44, 0·87) | 60·8 (44·5, 72·3) | 59·8 (47·2, 69·3) |
| Unvaccinated (N=50,831) | 42 | 0·083 (0·061, 0·11) | Ref. | Ref. | 803 | 1·58 (1·47, 1·69) | Ref. | Ref. |

Abbrevations, N: Number of women; CIN3+: cervical intraepithelial neoplasia grade 3 or worse; VE: vaccine effectiveness; Risk: Crude cumulative risk; CI: confidence interval; Ref.: reference category.
